# Psychiatric Genome-wide Association Study Enrichment Shows Promise for Future Psychopharmaceutical Discoveries

**DOI:** 10.1101/2023.12.05.23299434

**Authors:** Alexander S. Hatoum, Aaron Gorelik, Lauren Blaydon, Tingying Chi, David A.A. Baranger, Alex P. Miller, Emma C. Johnson, Arpana Agrawal, Ryan Bogdan

**Author notes:** Please send all correspondence to: Washington University School of Medicine, Department of Psychiatry, 660 S. Euclid, CB 8134, Saint Louis, MO 63110, USA. Disclosures:. Alexander S. Hatoum is list as an on inventors on two preliminary patents related to these analyses, T-020521 — “Treatments for multi-drug or broad addiction liability”, and T-020507 “Multi-omics algorithm for testing neurological and psychiatric pharmaceutical efficacy”.

## Abstract

Innovation in psychiatric therapeutics has stagnated on a handful of known mechanisms. Psychiatric genome-wide association studies (GWASs) have begun to identify hundreds of genome-wide significant (GWS) loci and characterize complex polygenic architectures. These data have rapidly advanced our understanding of disease etiology but their ability to inform clinical treatment in psychiatry remains largely unexplored. Here, we show that large psychiatric GWASs (schizophrenia, bipolar disorder, major depressive disorder, and substance use disorders) are enriched (OR: 2.776-27.629) for drug targets of current therapeutics used to treat these disorders (e.g., SNRI’s for MDD; antipsychotics for schizophrenia and MDD). The effect sizes of individual loci and the number of variants identified in a study, but not functional annotations of SNPs, drove enrichment. Psychiatric GWAS results may assist in drug repurposing and novel treatment identification.

## Main

Psychiatric disorders account for over 14% of deaths^1^ and 10% of lost years of health globally^2^. While preventative efforts and treatment developments have led to a reduction in the overall global burden of disease in recent decades^2^, the impact of psychiatric disorders has risen while drug development to treat these disorders has stagnated. Indeed, the unknown etiology of psychopathology has predominantly restricted medication development to a limited number of known pharmacologic drug targets (e.g., dopamine receptor 2 antagonists, selective serotonin/norepinephrine reuptake inhibitors) that were often discovered by happenstance (treatments being developed to treat tuberculosis and pain were discovered to have antidepressant and antipsychotic properties, respectively) and the reduction of side effects associated with these medications^3^. As major treatment advances often directly target links within an etiologic chain (e.g., statins inhibiting cholesterol synthesis), leveraging our developing understanding of psychopathology to guide treatment development holds promise.

Recent large scale psychiatric genome-wide association studies (GWASs) have begun to characterize their extensive polygenic architectures and reliably identified hundreds of common variants and pathways associated with risk^4^ that may be leveraged to prioritize novel drug targets and identify existing medications used to treat other conditions that may be repurposed to treat psychiatric disorders. Evidence supporting the promise of GWAS-informed treatment advancement comes from other fields. For example, early research across various categories of broad indications (e.g neurological disease, cardiovascular disease, etc.) found that medications acting on pathways implicated by GWAS are more likely to be approved therapeutics for diseases of that indication and progress further in clinical trials^5^. More recently, GWASs have contributed to novel U.S. Food and Drug Administration (FDA) approved treatments for COVID-19^6^. Whether GWAS data may be leveraged to hasten treatment development in psychiatry, where etiology remains poorly understood, remains unclear.

Here, as an initial proof-of-principal, we tested if therapeutics for specific psychiatric disorders are enriched for genome-wide significant signals within GWAS of that psychiatric disorder. We tested schizophrenia (SCZ; GWAS n=320,404), bipolar disorder (BiP; n=413,466); major depressive disorder (MDD; n=1,004,980), substance use disorders (SUDs; a general factor encompassing opioid use disorder, alcohol use disorder, tobacco use disorder, and cannabis use disorder; n=1,025,550), attention deficit hyperactivity disorder (ADHD; n=225,534), posttraumatic stress disorder (PTSD; n=174,659); generalized anxiety disorder (GAD; n=241,541), and insomnia (INSOM; n=386,533; Methods). We relied on proximity mapping (i.e., gene assignment based on the physical position of SNPs on the drug target), but also examined whether eQTL, chromatin refolding, genomic annotation or effect size further improves enrichment (**Methods**).

## Results

Proximity mapping revealed that targets of medications used to treat SCZ, BiP, MDD, and SUDs, but not ADHD, PTSD, GAD, or INSOM, are enriched for GWAS signals of their respective disorders, after Bonferroni correction for the number of disorders tested (odds ratios [ORs]: SCZ=27.629; BiP=5.398; MDD=2.776; SUDs=OR=11.95; all ps < 1.15e-3; **Fig. 1A**; **Supplemental Table 1; Methods**). Gene-drug-disease pairings (total n=74) included the following genes and medication classes (**Figure 1B**): **1) SCZ:** dopamine (e.g., *DRD2 antagonists*-antipsychotics, e.g., clozapine), glutamate (e.g., *GRM3, GRIN2A*-NMDA receptor modulators, e.g., haloperidol and risperidone); **2) BiP:** serotonin (e.g., *HTR6A*-antipsychotic class, e.g., quetiapine), and calcium channels (*CACNB2, CACNA1C, CACNA1B*-anticonvulsants, e.g., gabapentin); **3) MDD:** Norepinephrine reuptake drugs (specifically duloxetine, and Nortriptyline, with *NRXN1* and *NCAM1 involved in modulation of chemical synaptic transmission*); **4) SUDs:** dopamine (*DRD2*-dopamine agonists, e.g., bupropion), and substance-specific pathways (e.g. *CHRNB2-varenicline, OPRM1-naltrexone*). Our negative control analysis provided no evidence that psychiatric drug targets are enriched for type 2 diabetes genome-wide significant signals (P=.119-.993; **Methods; Supplemental Table 2**).

**Figure 1.**
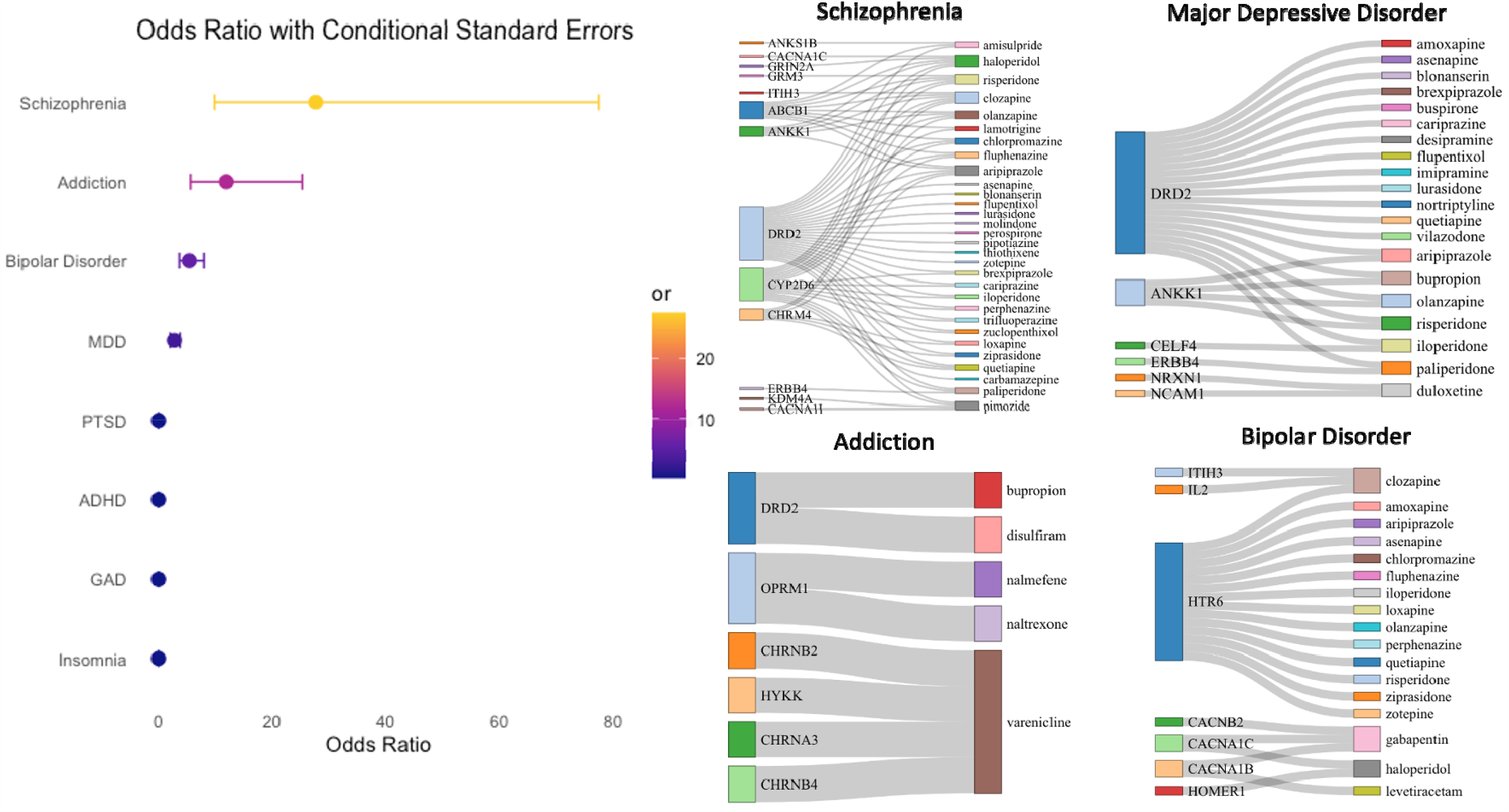
GWAS for psychiatric disorders are enriched for treatments of that disorder for the latest waves of Schizophrenia, SUDs/Addictions, Bipolar Disorder, and Major Depressive Disorder. **Note**. GWAS implicates current treatments for psychiatric disorders. **(A)** Odds ratio of a drug implicated by GWAS being a known treatment for a psychiatric disorder, controlling for the number of drug targets of each drug. Those odds ratios that were significant after Bonferroni are plotted with their confidence interval. MDD = Major depressive disorder, ADHD = Attention deficit hyperactivity disorder, GAD = generalized anxiety disorder, PTSD = Post-Traumatic Stress Disorder. **(B)** The link between a treatment for that disease and the drug targets from GWAS are shown as a Sankey Network Diagram. Larger colored edges signify genes (left) and drugs (right) that are more connected. These are the positive enrichment cases that are driving our significant findings.

Analyses accounting for shared targets across therapeutics (inclusion of medication class random intercepts; **Methods**) and alternative coding for drug targets (**Methods**) produced largely consistent results (**Supplemental Results, Supplemental Table 3**). For instance, *DRD2* GWAS signals drove enrichment for all schizophrenia targets; however, an analysis that excluded therapeutics that were only positive cases because of *DRD2* GWAS signals still revealed significant enrichment for SCZ treatments implicated by GWAS (OR=19.49; p=0.000247).

Next, we tested whether drug targets are enriched for genes identified through local expression quantitative trait loci (cis-eQTL) and conformational (i.e., chromatin interactions: Hi-C) annotation of GWAS signals, and whether such signals may supplement proximity matching drug target enrichment for GWAS signals (**Methods**). eQTL or conformational mapping (i.e., HiC) alone revealed less consistent therapeutic enrichment than proximity mapping (**Supplemental Table 4**). When cis-eQTL and Hi-C annotations were used to supplement proximity mapping, there was evidence for drug target enrichment; however, enrichment values were nominally lower and within the confidence intervals of proximity mapping alone (**Supplemental Table 5**).

Finally, we explored whether additional GWAS annotations (i.e., functional annotations including Combined Annotation Dependent Depletion [CADD] scores, regulomeDB scores, and gene region [e.g., exonic, intron]), or other information (i.e., effect size of genome-wide significant SNPs; or number of drug targets of that therapeutic implicated by GWAS; GWAS sample size) potentiated drug target enrichment for GWAS signals (see **Methods**). When annotating SNPs with additional information, the only consistent increase in enrichment (as indicated by non-overlapping confidence intervals) across all disorders was the largest effect size of any SNP, which increased enrichment betas by 18-44 fold (**Figure 2** and **Supplemental Table 7)**. GWAS sample size played a significant role in therapeutic enrichment; smaller (earlier) GWAS of SCZ, BiP, and MDD were not enriched for drug targets for therapeutics and enrichment was also correlated with the number of independent loci (r^2^ = .68; **Supplemental Results and Supplemental Figure 1; Supplemental Table 1**).

**Figure 2.**
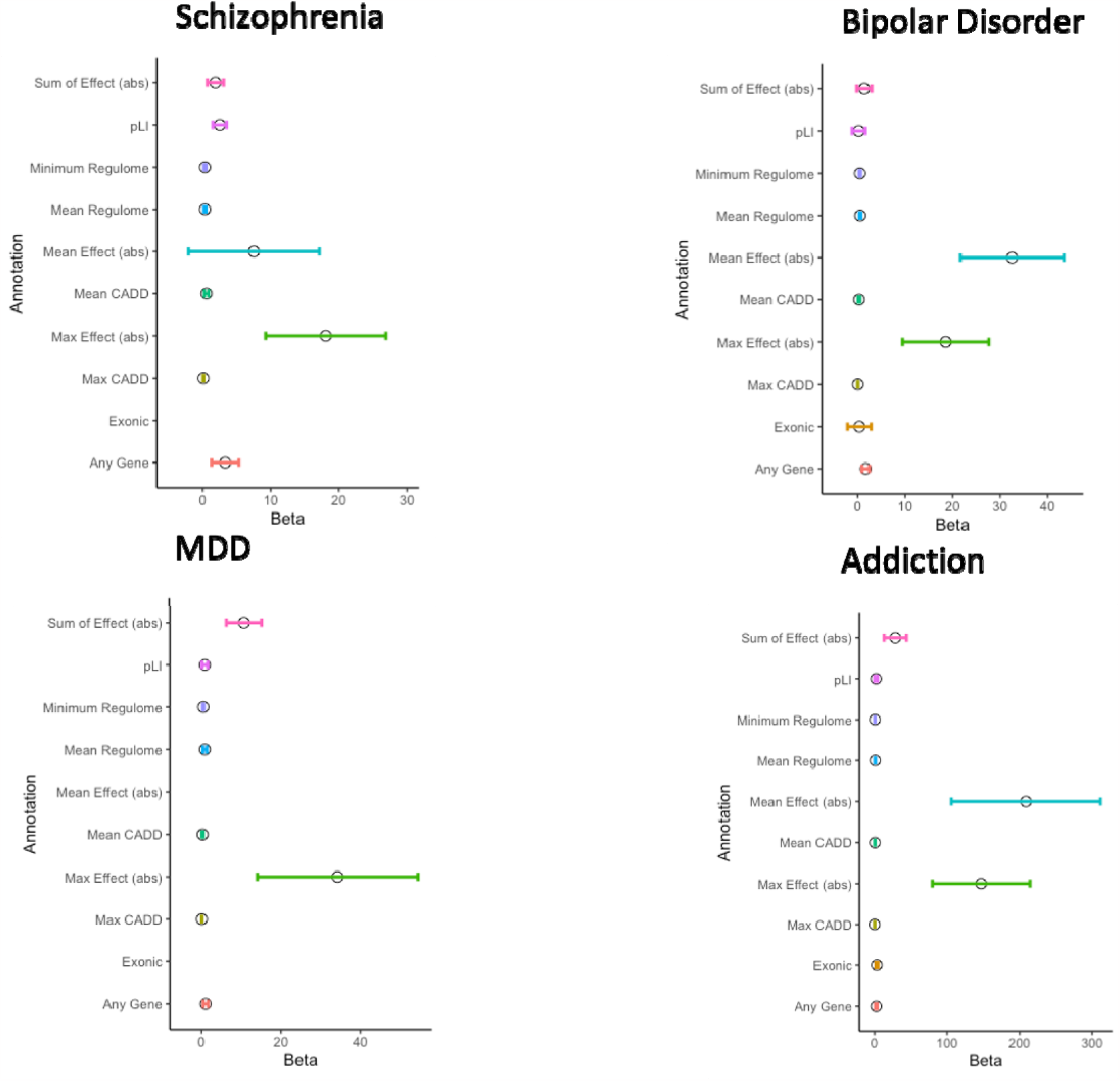
Enrichment for Various Genomic Annotations. For each disorder (MDD = major depressive disorder) we plot the enrichment when genomic annotations are used instead of 0 or 1 any-gene pairing. Each dot is the beta and confidence intervals for that beta. Abs = absolute value or that the absolute value of SNP effect sizes was used, CADD = Combined Annotation Dependent Depletion, pLI = Loss Intolerance probability score. When points are missing from the plot, it is because no Exonic variant was a drug target after pruning SNPs for linkage disequilibrium.

## Discussion

The multitude of mechanisms contributing to the expression of psychiatric disorders has made it challenging to identify biological pathways that can be therapeutically leveraged^7^. Indeed, recent treatment developments in psychiatry have been largely constrained to refinements of existing treatments that were developed decades ago^3^. Our findings that current psychiatric disorder drug targets are enriched for genes identified through GWAS provides initial evidence that GWAS may potentially identify and prioritize treatment targets for drug development and identify existing drugs that could be repurposed and/or reformulated to reduce the burden of psychiatric disorders. Proximity matching largely drove enrichment with limited benefit of additional annotation (aside from effect size); as drug enrichment was greater in larger GWASs, these results highlight the potential clinical importance of further expanding GWAS samples.

While our data showcase the potential of GWASs to enhance and expedite the treatment of psychiatric disorders, challenges plaguing psychiatric treatment remain. Disorders represent heterogeneous amalgamations of symptoms with the same diagnosis having many distinct putative etiologies and comorbidity being commonplace. Advances in nosology may facilitate a better understanding of shared and distinct etiologies to facilitate the isolation of more specific, and potentially less generalized, therapeutic pathways^8^.

In addition to these broad psychiatric treatment challenges, the use of GWAS data to inform therapeutic development has several limitations. First, acute transcription changes, etc. May not identify current treatment downstream mechanisms, which may be particularly relevant for treatments that do not show benefit until later - e.g., SSRI and trkb, etc^9^. Second, GWASs have largely been restricted to European ancestry with little representation from diverse global populations^10^. Third, there are large sex differences in psychopathology, yet GWASs may not inform sex-specific treatments as sex stratified analyses and evaluation of sex chromosomes are rare in psychiatric GWAS. Third, relying on GWASs for drug development prioritizes common variability in the genome; rare variants have been studied in schizophrenia and cluster in similar areas of the genome as GWAS hits, but with larger effect sizes, suggesting potential for drug repurposing based on current analyses^11^. Finally, many loci associated with disorders through GWAS represent broad regulators of multiple pathways (e.g., *PDE4*^12^) that may be difficult to target without inducing side effects and further research may be needed to identify which affected pathways may have etiologic significance.

Despite limitations, GWAS data validate current drug targets for psychiatric medications. This proof of principle highlights the clinical potential of psychiatric GWASs to expand treatment by identifying novel treatment targets and existing drugs for other conditions that could be repurposed. Ultimately, the ability to validate discoveries from biological psychiatry may improve innovation and reduce the disproportionate rising burden of mental illness.

## Online Methods

### Discovery GWAS

Summary statistics from the latest genome-wide association studies of psychiatric disorders with at least 3 independent genome-wide significant loci were included: **1)** schizophrenia N = 320,404 (Wave 3; Trubetskoy, Pardinas, et al., 2022^13^), **2)** bipolar disorder (BiP) N = 413,466 (Wave 3; Mullins, Forstner, et al., 2021^14^), **3)** major depressive disorder (MDD) N = 1,004,980 (Levey et al., 2021^15^), **4)** generalized anxiety disorder (GAD), N=241,541 (Levey et al., 2020^16^), **5)** attention deficit hyperactivity disorder (ADHD) N=225,534 (Demontis et al., 2022^17^), **6)** posttraumatic stress disorder (PTSD) N = 174,659 (Stein et al., 2019^18^), **7)** general and specific substance use disorders, N=1,025,550 (Hatoum et al., 2023^19,20^). Insomnia (N = 386,533; Jansen et al., 2019^21^) was also included as it is a transdiagnostic feature of many psychiatric disorders and has dedicated treatments, many of which (e.g., quetiapine, trazodone) are also used to treat psychiatric disorders. Eating disorders were excluded due to the lack of known treatments. Finally, as a negative control we used a GWAS of diabetes from the DIAGRAM consortium, n = 1,339,889 (Mahajan et al., 2022^22^). All summary statistics retained only common SNPs with Minor Allele Frequency (MAF) ≥ .01 and were aligned to genome build 37.

To enable tests of whether GWAS power/sample size impacts drug identification, we also used earlier GWAS waves of these same psychiatric disorders, which included: **1)** schizophrenia Psychiatric Genomics Consortium (PGC) wave 1 (N =51,695)^23^, **2)** schizophrenia PGC wave 2 (N = 150,064)^24^, **3)** bipolar disorder wave 1 (N = 16,731)^25^, **4)** bipolar disorder wave 2 (N = 51,710)^26^, and **5)** major depressive disorder (N= 480,359)^27^.

### SNP-Gene Pairing

#### Proximity Matching

FUMA^28^ was used to assign independent (r^2^ < .6) genome-wide significant SNPs identified in GWASs to genes ± 10 kbp (excluding the MHC region due to its complex long range linkage disequilibrium) based on SNP physical location. All GWASs were aligned to human genome build 37.

### Drug-Psychiatric Disorder Pairing

The *Prescriber’s Guide: Stahl’s Essential Psychopharmacology version 7* (Stahl’s guide)^29^ was used to identify drugs to treat: ADHD (n treatments=18), MDD/depression (n treatments=51), schizophrenia/psychosis (n=31), bipolar/mania/maintenance (n treatments=25), PTSD (n treatments=25), GAD (n treatments=35), and insomnia (n treatments=15). Due to the small number of treatments per substance use disorder and the shared genetic etiology^20^, substance use disorders were combined into a single category (alcohol use disorder/dependence, opioid use disorder, tobacco/nicotine dependence, cannabis use disorder; n treatments=8). This also allowed us to match to the largest study of generalized genetic risk for SUDs to date (Hatoum et al. 2023^19^). We chose Stahl’s guide for our primary analysis because the guide includes a combination of FDA approved therapeutics, along with commonly used therapeutics that are prescribed off label, and adjuncts. This gives us a more dense list of drugs that are likely to be true positives than relying only on the U.S. Food and Drug Administration approval. Double data entry was used to ensure accuracy of the coding scheme.

#### Defining Drug Targets

Gene-Drug pairings were defined using the Drug Gene Interaction Database (DGldb; which uses a dense literature search to assign drug targets^30^) and Connectivity Map (CMAP; which uses differential expression from human cell lines^31^), both of which have been previously used to study psychiatric drug repurposing (N_Drug_ = 1,201^32^).

#### Alternative Coding Schemes

To test for robustness across coding schemes for drug-disease and gene-drug pairing, we considered an alternative gene-drug pairing coding scheme using OpenTargets.org^33^ and an alternative drug-disease pairing scheme using clinicaltrials.gov. Open Target Genetics is an online tool and database which integrates functional and biological information from numerous sources including the literature and other large open science studies (i.e., UK-Biobank) to enable identification of potential drug targets. For each drug we extracted a complete list of drug targets and their proximal genes. We labeled a drug as missing if the drug either did not have any potential drug targets or was not listed in the database. We used double data entry to ensure accuracy of this coding scheme.

As an alternative dataset of drug-disease pairing, we used clinicaltrials.gov, an open-source repository of clinical research studies (i.e., clinical trials and observational studies). Importantly, this database is reliant on both sponsors and/or investigators to submit records of their studies and only a limited review by the national library of medicine is conducted. The following inclusion criteria were used: 1) clinical trials; 2) interventional studies; and 3) trials phase (one through four). Trials with results and the following information were extracted from each drug for the highest clinical trial phase of each psychiatric disorder: 1) number of highest trial phase completed, 2) clinical trial status (e.g. if terminated, why); 3) type of result (i.e.., positive or negative); and 4) any additional notes to be reviewed by the rest of the coding team. We had a 4-level coding scheme for drug-disease pairing evidence. The highest degree of clinical trial success that is currently known (i.e., 1, 2, 3, 4), or if the drug is FDA approved but not on clinicaltrials.gov, it was given a 4. If it was in Stahl’s but not on ClinicalTrials.gov, it was given a 3. The variable was treated as a 4-level variable in a logistic regression to test for enrichment for each respective drug (see model below). This was to mirror past analyses that compared enrichment across stages of clinical development (see Nelson et al., 2015^5^). We used double data entry to ensure accuracy of this coding scheme.

### Disorder Gene-Drug Pairing Enrichment Analyses

We tested if drug targets (e.g., transcripts or proteins directly modulated by the drug) of therapeutics are enriched for GWAS signals associated with the psychiatric conditions they are used to treat. In other words, a drug-gene pairing was considered a positive enrichment case when a SNP reaching genome-wide significance in a GWAS of that disorder resides within a gene (± 10 kbp) coding for a protein that is targeted by a therapeutic approved to treat that disorder.

> Logistic regression models were implemented in base R (version 4.2.0)^34^ as follows:
>
> Y = *β*_0_ + X_1_*β*_1_ + X_2_*β*_2_
>
> Y = 0 or 1 if therapeutic treats the specific psychiatric disorder.
>
> *β*_0_ = Model intercept.

X_1_ = Number of therapeutic drug targets (positive whole number integer) was included as a covariate. Because targeting expression of many genes could lead to a therapeutic being more likely to be a treatment and also more likely to contain a gene implicated by GWAS, we found this covariate to be a necessary baseline in order to appropriately evaluate our findings. In other words, our test is a competitive test, with the baseline for enrichment being having more drug targets.

X_2_ = 0 or 1 based on if any gene implicated by the GWAS is a target for the psychiatric disorder (i.e., 0 if no drug targets of that therapeutic were found in the GWAS, 1 if any drug target of that therapeutic is mapped to the GWAS).

*β*_2_ is the primary parameter of interest, testing the null hypothesis of *β*_2_ = 0 against the alternative *β*_2_ ≠ 0, with a significant test indicating greater likelihood that a therapeutic with a drug target implicated by the GWAS is a current treatment used for that psychiatric disorder. As the outcomes are dichotomous values of overlap or no overlap, the test can be interpreted as odds increase in the chance that a drug paired with a disease by GWAS is a drug that is known to treat that psychiatric disorder, accounting for the number of drug targets of each therapeutic.

To test whether drugs with more drug targets that were implicated by GWAS were more enriched for current treatments, we also tested a model where X_2_ = a positive integer of the total number of drug targets implicated by the GWAS of that disorder.

Sankey diagrams were used to visualize drug-gene pairings using the network3D package in R^35^.

#### Alternatives to proximity mapping

##### eQTL and Hi-C

Next, as an alternative coding scheme to genes in proximity, we remapped genes with two additional criteria. Specifically, we tested *cis*-eQTL and HiC chromatin loops to see if these biological characterizations improved enrichment on their own, or when combined with proximity matching. Both used PsychENCODE^36^ data to assign SNP annotations, with Hi-C using the PsychENCODE EP links and PsychENCODE Promotor Anchored loops specifically. eQTLs were considered significant at fdr corrected *p <.05* in PsychENCODE.

#### Functional Annotation and Effect Size to Improve Enrichment

We added information based on annotation categories and effect size of SNPs in the GWAS to see if this information increased drug enrichment for genes associated with the disorder through GWAS. Each category was tested independently for enrichment. Functional Gene Annotation was done using the SNP2Gene function on FUMA (see: https://fuma.ctglab.nl/tutorial#snp2gene^10^). Specifically, we examined whether the variant that implicated the gene paired to the therapeutic was exonic based on ANNOVAR^37^ and whether the variant that implicated the gene was a protein truncating loci (pLI) based on the ExAC database^38^. Next, we developed several measures by using existing SNP functional annotation scores. First, we used the Combined Annotation Dependent Depletion (CADD) score^39^, a score that captures the expected deleteriousness of a SNP. We annotated therapeutics for the maximum CADD score of any SNP on the drug target for that therapeutic and the average CADD scores of all SNPs on drug targets for that therapeutic. We also used regulomeDB scoring^40^, which gives a rank to non-coding variants based on their likely regulatory role, with lower values showing increased likelihood of a regulatory effect. We annotated therapeutics for minimum regulomeDB score number and average regulomeDB score. Finally, we annotated therapeutics with several values representing the effect size of SNPs on the drug target. This included the highest effect size of a SNP on any gene that is a drug target for that therapeutic, mean effect size of a SNPs on drug targets for therapeutic, and sum of the absolute value of effect sizes of SNPs on the drug targets for that therapeutic.

## Data availability

The MVP summary statistics were obtained via an approved dbGaP application (phs001672.v4.p1). For details on the MVP, see https://www.research.va.gov/mvp/ and Gaziano, J.M. et al. Million Veteran Program: A mega-biobank to study genetic influences on health and disease. J Clin Epidemiol 70, 214-23 (2016)). This research is based on data from the Million Veteran Program, Office of Research and Development, Veterans Health Administration, and was supported by the Veterans Administration (VA) Cooperative Studies Program (CSP) award #G002.

All other data was downloaded through the Psychiatric Genomics Consortium Website (see: https://pgc.unc.edu/for-researchers/download-results/) and is freely available.

FUMA is an online platform and all data used are available through that platform: https://fuma.ctglab.nl/

All code is available through the first authors github: https://github.com/AlexHatoum

## Acknowledgements

The Authors would like to thank Cold Harbor Laboratory for posting a preprint of this work on MedArchive. The authors thank Million Veteran Program (MVP) staff, researchers, and volunteers, who have contributed to MVP, and especially participants who previously served their country in the military and now generously agreed to enroll in the study. Finally, the authors would like to acknowledge the Psychiatric Genomics Consortium for their open data policy.

## Author Contributions

ASH wrote the manuscript, designed pipelines for analysis, coded data, and conducted all analysis. AG and LB conducted data coding and conducted double data entry. TC provided clinical perspective on findings and reevaluated coding schemes to ensure they met expectations of a practicing community clinician. The analysis was conducted in the lab of RB who provided critical organization and commentary on the manuscript and all analyses. All authors (including AA, APM, DAAB, and ECJ) gave critical feedback on the manuscript and analyses.

## Inclusion & Ethics in Global Research Statement

No data or research was provided by individuals from the global south. This paper meets the standards of Inclusion & Ethics in Global Research required by Springer.

