## Supplemental Tables and Figures for "Psychiatric Genome-wide Association Study Enrichment Shows Promise for Future Psychopharmaceutical Discoveries"

**Supplemental Materials**

**Index**

Pg. 1 Supplemental Results

**Supplemental Tables**

Pg. 2  Supplemental Table 1:Enrichment Values By Each Disease Test for Treatments of That Disease
Pg. 3. Supplemental Table 2:Enrichment of a Diabetes GWAS for Psychopharmaceuticals as a Negative Control
Pg. 4 Supplemental Table 3: Enrichment in a mixed effects model with random intercepts for class or Super Class
Pg. 5 Supplemental Table 4: Enrichment for Significant Disease-Drug-Gene traits by Alternative Coding Schemes
Pg 6. Supplemental Table 5: Enrichment Using Cis-eQTL or HiC to Assign Mapped Genes Instead of Closest Gene
Pg. 7. Supplemental Table 6:  Enrichment Supplementing Proximity Mapping with Cis-eQTL and Hi-C
Pg. 8. Supplemental Table 7:  Enrichment of Psychopharmaceuticals by Various Genomic Annotations

**Supplemental Figures**

Pg. 10.  Supplemental Figure 1: Sample Size Dependency of Enrichment

### Supplemental Results

*Alternative Coding Scheme Results*. We reran enrichment analyses using two alternative coding schemes in order to evaluate robustness. As an alternative coding scheme to Stahl's prescriber guide to assign drug-disease pairing, we added information on clinical trial stage completed using clinicaltrials.gov. As an alternative to the gene-drug pairing using cMAP and Drug Interaction Database we used OpenTargets.org. Our results were largely consistent with the main results.

Using clinicaltrials.gov to assign alternative drug-disease indications yielded significant results for bipolar disorder, schizophrenia, and major depressive disorder **(Supplemental Table 4**). Substance use disorders could not be tested because the combined category was not possible as many drugs that work for one disorder are tested and failed for others at different clinical trial stages.

We used an alternative gene-drug pairing using OpenTargets.org. We found significant enrichment for Schizophrenia, Major Depressive disorder, and SUDs. We did not find significant enrichment for bipolar disorder. Upon closer inspection, for most psychiatric medications OpenTargets.org contains more restricted lists of gene-drug targets that are only the known functional target that is thought to produce the effect. This means that serotonergic targeting drugs only listed *SLC6A4* or *SLC6A2*, and typically just *SLC6A4*. This left out their known receptors, which drove many results in bipolar disorder in the main analysis (namely *HTR6A*). We concluded through exploration that these lists were likely overly restrictive. For example, Nortriptyline does not list *DRD2* or SNRI targeting mechanisms, but is a well-known modulator of norepinephrine and a suspected modulator of dopamine, in addition to the main mechanism of action. In general, Tricyclic antidepressant and anti-convulsants did not list off-targets, but these off-targets are often the mechanism of effectiveness for the "off-label" prescribed medications commonly used to treat bipolar disorder (for example *CACNA1C*). The assumption that we should limit to the main targets of the mechanism of a drug is also restrictive for future drug repurposing efforts, as off-targets may produce therapeutic effects that are desired. Notably, despite the conservative list, 3 of 4 traits remained significant using OpenTargets.org

*GWAS discovery power is likely tied to enrichment.* We conducted empirical analysis of the degree to which earlier smaller GWASs of SCZ, BiP, and MDD were enriched for psychopharmaceuticals, with smaller waves typically not enriched (**Supplemental Table S1**); 2) all GWAS with significant treatment enrichment identified more genome-wide significant loci than those that did not (i.e.,for MDD,Bip, SCZ mean N hits = 310 with a range of 86-656, vs N hits = 23 and a range of 4 to 77 for non-significant findings; **Supplemental Table 1** and **Supplemental Figure S1**), and 3) the number of discovered loci increases the odds of enrichment (**Supplemental Figure S1**; r^2 = .6 between enrichment effect size and independent significant hits found).

### Supplemental Tables

**Supplemental Table 1**. Enrichment Values By Each Disease Test for Treatments of That Disease (Visualized in Figure 1)

| Trait | Independent Hits | Mapped Genes | Log(Odds) | SE | P-value |
|---|---|---|---|---|---|
| **SCZ1** | 12 | 77 | -15.800799 | 799.240094 | 9.84E-01 |
| **SCZ2** | 352 | 452 | 3.704155 | 1.025073 | 0.000302 |
| **SCZ3** | 656 | 713 | 3.318882 | 1.026613 | 0.00123 |
| **MDD1** | 16 | 96 | -12.271863 | 727.202703 | 0.986536 |
| **MDD2** | 295 | 303 | 1.02112 | 0.314147 | 0.00115 |
| **BIP1** | 10 | 8 | -13.578407 | 771.499873 | 0.986 |
| **BIP2** | 23 | 64 | 0.030375 | 0.670221 | 0.964 |
| **BIP3** | 86 | 259 | 1.686041 | 0.394786 | 1.95E-05 |
| **PTSD** | 4 | 5 | -13.181676 | 882.743585 | 0.988 |
| **ADHD** | 77 | 61 | -14.837923 | 964.757768 | 0.988 |
| **SUDs** | 163 | 146 | 2.48076 | 0.75338 | 0.000992 |
| **GAD** | 7 | 6 | -15.559783 | 787.28408 | 0.98423 |
| **Insomnia** | 35 | 25 | -15.68601 | 1295.00539 | 0.9903 |

Note. Table of enrichment values across all traits studied. SE = standard error Schizophrenia 1 = Schizophrenia Wave 1, SCZ2 = Schizophrenia Wave 2, SCZ3 = Schizophrenia Wave 3 (main analysis), MDD1 = Major Depressive Disorder Wave 1, MDD2 = Major Depressive Disorder Wave 2 (main analysis), BiP 1 = Bipolar Disorder Wave 1, BIP 2= Bipolar Disorder Wave 2, BIP 3 Bipolar Disorder Wave 3 (main analysis), PTSD = Post-Traumatic Stress Disorder, ADHD = Attention Deficit Hyperactivity Disorder, GAD = Generalized Anxiety Disorder.

**Supplemental Table 2**. Enrichment of a Diabetes GWAS for Psychopharmaceuticals as a Negative Control

| **Diabetes Enrichment** | **Beta** | **SE** | **P** |
|---|---|---|---|
| **MDD** | -0.715 | 0.459 | 0.119352 |
| **SCZ** | 0.0043 | 0.499 | 0.993 |
| **BiP** | -0.254 | 0.486 | 0.601 |
| **SUDs** | -1.513 | 1.445 | 0.18 |

Note. MDD = Major Depressive Disorder, SCZ = schizophrenia, BiP = Bipolar disorder.

**Supplemental Table 3**. Enrichment in a mixed effects model with random intercepts for class or Super Class

| **Trait** | **Beta** | **SE** | **P** |
|---|---|---|---|
| **Class** | | | |
| **MDD** | 1.0092 | 0.3807 | 0.00802 |
| **SCZ** | 2.68113 | 1.06402 | 1.17E-02 |
| **BiP** | 2.02914 | 0.56316 | 0.000314 |
| **SUD** | 2.479771 | 0.75342 | 0.000997 |
| **Super Class** | | | |
| **MDD** | 1.072497 | 0.3195 | 0.000789 |
| **SCZ** | 3.192711 | 1.030163 | 0.00194 |
| **BiP** | 1.54884 | 0.40171 | 0.000115 |
| **SUD** | 2.52565 | 0.75755 | 0.000856 |

Note. Enrichment values when modeling the enrichment as a logistic mixed effects model with a random intercept for class or Super Class as determined by ClassifyFire chemical analysis.

**Supplemental Table 4**. Enrichment for Significant Disease-Drug-Gene traits by Alternative Coding Schemes

| OpenTargets.org Enrichment | Beta | SE | P |
|---|---|---|---|
| MDD | 2.088 | 0.37 | 1.71E-08 |
| SCZ | 3.041 | 0.4 | 3.07E-14 |
| BIP | 0.327 | 0.807 | 0.6448 |
| SUDs | 2.135 | 0.759 | 0.00488 |

| ClinicalTrials.gov enrichment | Beta | SE | P |
|---|---|---|---|
| MDD | 0.953 | 0.306 | 0.00182 |
| SCZ | 1.47 | 0.39 | 1.61E-04 |
| BIP | 1.12 | 0.362 | 0.00205 |

Note. Alternative coding scheme enrichment for opentargets.org to define drug-gene pairing and clinicalTrials.gov to define disease drug-pairing. SUDs could not be tested with clinicaltrials.gov since most clinical trials are for specific SUDs and restricted to single substances. Because of this, coding to resolve the degree of completed trial for a treatment would only be able to use information for a single substance use disorder trait at time. MDD = Major Depressive Disorder, SCZ = schizophrenia, BiP = Bipolar disorder.

**Supplemental Table 5**. Enrichment Using Cis-eQTL or HiC to Assign Mapped Genes Instead of Closest Gene

| **cis-eQTL enrichment** | **Beta** | **SE** | **P** |
|---|---|---|---|
| **MDD** | 0.166 | 0.554 | 7.64E-01 |
| **SCZ** | 0.544 | 0.41 | 1.85E-01 |
| **BiP** | 1.686 | 0.395 | 1.95E-05 |
| **SUDs** | 0.503 | 0.594 | 0.397 |

| **HiC Enrichment** | **Beta** | **SE** | **P** |
|---|---|---|---|
| **MDD** | 1.441 | 0.313 | 4.26E-06 |
| **SCZ** | 4.058 | 0.741 | 4.28E-08 |
| **BiP** | -0.319 | 0.689 | 0.643 |
| **SUDs** | 1.41 | 0.761 | 0.0642 |

Note. MDD = Major Depressive Disorder, SCZ  = schizophrenia, BiP = Bipolar disorder.

**Supplemental Table 6**. Enrichment using any gene that mapped to eQTL, Hi-C or through proximity mapping

| **Trait** | **Beta** | **SE** | **P** |
|---|---|---|---|
| **SCZ** | 3.199 | 1.027 | 0.00185 |
| **BiP** | 1.431 | 0.409 | 0.000464 |
| **MDD** | 0.7801 | 0.315 | 0.0134 |
| **SUDs** | 1.286 | 0.311 | 3.48-e05 |

Note. MDD = Major Depressive Disorder, SCZ  = schizophrenia, BiP = Bipolar disorder.

**Supplemental Table 7**. Enrichment of Psychopharmaceuticals by Various Genomic Annotations

| | Schizophrenia | | | MDD | | |
|---|---|---|---|---|---|---|
| | Enrichment Beta | SE | P | Enrichment Beta | SE | P |
| **Any Gene** | 3.318 | 1.02 | 0.0012 | 1.021 | 0.3141 | 0.0012 |
| **Sum of Effect (abs)** | 1.882 | 0.6094 | 0.002 | 10.661 | 2.259 | 2.37E-06 |
| **Max Effect (abs)** | 18.081 | 4.5052 | 5.98E-05 | 34.263 | 10.324 | 0.0009 |
| **Mean Effect (abs)** | 7.533 | 4.902 | 0.124 | -27.204 | 19.3161 | 0.159 |
| **Max CADD** | 0.113 | 0.027 | 3.12E-05 | 0.048 | 0.0152 | 0.0016 |
| **Mean CADD** | 0.566 | 0.156 | 0.000287 | 0.249 | 0.0899 | 0.0055 |
| **Minimum Regulome** | 0.346 | 0.0812 | 2.09E-05 | 0.4285 | 0.0816 | 1.51E-07 |
| **Mean Regulome** | 0.346 | 0.0812 | 2.09E-05 | 0.819 | 0.2468 | 0.0009 |
| **Exonic** | -16.259 | 946.681 | 9.86E-01 | NA | NA | NA |
| **pLI** | 2.521 | 0.5143 | 9.49E-07 | 0.825 | 0.3774 | 0.0289 |
| | **Bipolar Disorder** | | | **SUDs** | | |
| **Any Gene** | 1.686 | 0.3948 | 1.95E-05 | 2.48 | 0.7534 | 0.001 |
| **Sum of Effect (abs)** | 1.397 | 0.8042 | 8.23E-02 | 28.724 | 7.86 | 0.0003 |
| **Max Effect (abs)** | 18.546 | 4.633 | 6.26E-05 | 147.07633 | 34.3351 | 1.85E-05 |
| **Mean Effect (abs)** | 32.518 | 5.553 | 4.76E-09 | 208.786 | 52.4009 | 6.77E-05 |
| **Max CADD** | 0.046 | 0.0231 | 4.52E-02 | 0.1356 | 0.03633 | 0.0002 |
| **Mean CADD** | 0.301 | 0.1079 | 5.30E-03 | 0.6614 | 0.2079 | 0.0015 |
| **Minimum Regulome** | 0.47 | 0.0774 | 1.21E-09 | 0.6894 | 0.169 | 4.51E-05 |

| | | | | | | |
|---|---|---|---|---|---|---|
| **Mean Regulome** | 0.522 | 0.0812 | 1.33E-10 | 1.055 | 0.263 | 6.04E-05 |
| **Exonic** | 0.3599 | 1.269 | 0.777 | 3.37 | 0.7781 | 1.52E-05 |
| **pLI** | 0.2255 | 0.7066 | 7.50E-01 | 2.22 | 1.0374 | 3.24E-02 |

Note. Enrichment for targets of psychopharmaceuticals when using genomic annotations and values of genomic annotations. pLI = protein truncating loci (0, 1), abs = absolute value, CADD = Combined Annotation Dependent Depletion. Visuzlied in Supplemental Figure 2.

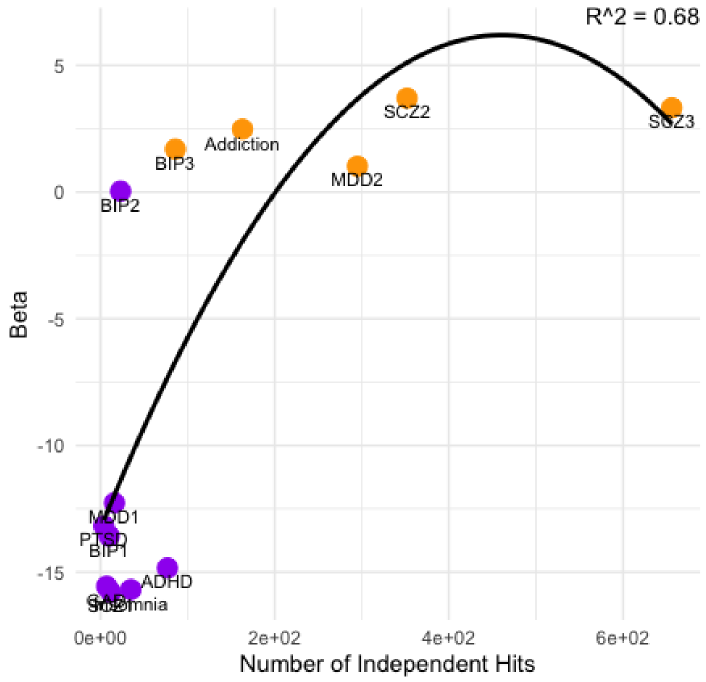


**Supplemental Figure 1. Effect size of enrichment plotted against number of independent loci shows that enrichment of GWAS for current treatments is highly associated with the number of independent loci discovered by that GWAS.** Beta representing an enrichment value is plotted on the y axis against the number of independent hits (SNPs with p-values less than 5e-8 that are greater than r^2 = .68 distance from each other). Each trait studied is noted by a single dot and a quadratic line is fit to the effect. Those traits that were significant are colored orange, those that are non-significant are colored purple. The figure shows there is a large and apparent effect of the number of GWAS independence loci discovered on enrichment of that GWAS for current treatments for that disorder. MDD2 = Major depression disorder wave 2, MDD1 = Major depression Disorder Wave 1, BIP1 = Bipolar Disorder Wave 1, BIP2 = Bipolar Disorder Wave 2, BIP3 = Bipolar Disorder Wave 3, SCZ 1 = Schizophrenia Wave 1, SCZ2 = Schizophrenia Wave 2, SCZ3 = Schizophrenia Wave 3, ADHD = Attention deficit hyperactivity disorder, GAD = generalized anxiety disorder, PTSD = Post-Traumatic Stress Disorder. Insomnia and SUDs are also shown.
